# Exploring the perceptions and experiences of community rehabilitation for Long COVID from the perspectives of Scottish General Practitioners’ and people living with Long COVID: a qualitative study

**DOI:** 10.1101/2023.11.06.23298096

**Authors:** Kay Cooper, Edward Duncan, Erin Hart-Winks, Julie Cowie, Joanna Shim, Emma Stage, Tricia Tooman, Lyndsay Alexander, Alison Love, Jacqui Morris, Jane Ormerod, Jenny Preston, Paul Alan Swinton

## Abstract

**Objectives:** To explore the experience of accessing Long COVID community rehabilitation from the perspectives of people with Long COVID and General Practitioners (GPs).

**Design:** Qualitative descriptive study employing one-to-one semi-structured virtual interviews analysed using the framework method.

**Setting:** Four NHS Scotland territorial health boards.

**Participants:** Eleven people with Long COVID (1 male, 10 female; aged 40-65 [mean 53], and 13 GPs (5 male, 8 female).

**Results:** Four key themes were identified: i) The lived experience of Long COVID; ii) The challenges of an emergent and complex chronic condition; iii) Systemic challenges for Long COVID service delivery, and iv) Perceptions and experiences of Long COVID and its management, including rehabilitation.

**Conclusions:** There are several patient, GP, and service-level barriers to accessing community rehabilitation for Long COVID. There is a need for greater understanding by the public, GPs, and other potential referrers of the role of community rehabilitation professionals in the management of Long COVID. There is also a need for community rehabilitation services to be well promoted and accessible to the people with Long COVID for whom they may be appropriate. Service providers need to consider availability and accessibility of Long COVID rehabilitation and ensure adequate interprofessional communication and collaboration to enhance the experience for people with Long COVID.

**Strengths and limitations of this study:**

- This is the first study to explore the issue of accessing Long COVID community rehabilitation from the perspectives of potential service users and referrers in the Scottish context.
- One researcher conducted all interviews, ensuring consistency in their conduct
- Data were analysed and interpreted by multiple researchers, including people with Long COVID
- The small sample size, largely drawn from health boards with a similar approach to Long COVID rehabilitation, limits generalisability

## INTRODUCTION

Long COVID in adults is a multisystemic condition described as signs and symptoms that develop during or after an infection consistent with COVID-19, and continue for more than 12-weeks, which cannot be explained by an alternative diagnosis [1]. While global prevalence remains unclear, the World Health Organisation estimated that as many as 34 million people may have experienced Long COVID by 2022. [2] In the UK, the Office for National Statistics estimated that 1.9 million people (2.9% of the UK population) had self-reported Long COVID as of March 2023 [3]. The effect of Long COVID on individuals varies. More than 200 symptoms have been identified affecting multiple organ systems including thrombotic and cerebrovascular disease, chronic fatigue, and dysautonomia, resulting in significant reduction in quality of life [4].

Healthcare professionals, and general practitioners (GPs) in particular, are faced with complex challenges associated with the diagnosis and management of people with Long COVID (PwLC) as they attempt to disentangle nonspecific symptomology associated with the virus from those related to other conditions [5]. The National Institute for Health and Care Excellence (NICE) rapid guideline on Long-term effects of COVID-19 recommends that individuals have access to rehabilitation that is personalised and multi-disciplinary in nature [1]. Subsequent studies have emphasised the importance of this approach [6–8]. However, uncertainty remains around the effectiveness of rehabilitation, with research ongoing [9 10]. In England, service provision for Long COVID has centred around the development of specialised clinics, usually comprising multidisciplinary teams that include medical, allied health and psychology professionals[11]. In Scotland, Long COVID rehabilitation is delivered in community (non-hospital) settings by generalist allied health professionals, with local variation in models of service delivery [12].

PwLC have previously reported barriers to accessing healthcare and difficulties in navigating interactions with disjointed healthcare services [12–15]. Kingstone et al. [16] highlighted the importance of “finding the right GP” for PwLC accessing primary care. GPs are the first point of contact for PwLC and have an important role in ensuring that people receive appropriate treatment and are referred to specialist services, including community rehabilitation, when needed. GPs, however, continue to face uncertaintly around providing a Long COVID diagnosis and ongoing management and, furthermore, experience challenges referring PwLC onto appropriate diagnostic and interventional services [15 17].

There is a growing body of research on the lived experience of PwLC [18 19] including their self-reported barriers to accessing healthcare [13 14]. Until recently, however [15], none have focussed on community rehabilitation. There is also a growing body of research on the role of the GP in providing care to PwLC, including the challenges of diagnosis and medical management [17 20]. Brennan et al’s scoping review [17] included 19 studies on the management of Long COVID by GPs. They reported five articles that described difficulties with accessing services, particularly those in the communtiy such as follow-up services and Long COVID clinics [17]. Bachmeier et al’s qualitative study found that German GPs rarely referred their Long COVID patients to specialist care or rehabilitation [20]. This was partly due to referral restrictions in the German healthcare system and lengthy waiting times for rehabilitation services or post-COVID clinics.

The current study explored PwLC and GPs’ perspectives of accessing Long COVID community rehabilitation in the context of four Scottish regional health boards. Rehabilitation in this study is defined as intervention/s aimed at optimising function and reducing disability [21], delivered by any appropriate healthcare professional in a community setting. Community rehabilitation typically takes place in clinics and people’s homes. This work was embedded within a larger realist evaluation study called LOCO-RISE, investigating models of service delivery for Long COVID community rehabilitation in Scotland. The initial phase of the realist evaluation (November 2021 – April 2022) focussed on PwLC and service providers’ perspectives, experiences, and outcomes of Long COVID rehabilitation [15]. During this time period, we observed that in some areas, there were low numbers of PwLC receiving community rehabilitation services [15]. We sought approval for an amendment to the study (Ref: 21/WA/0118 A03) to understand potential reasons for these low numbers, in the context of the increasing number of PwLC, recommendations for providing community rehabilitation to PwLC [6 8 22] and knowledge that services existed across Scotland [12]. The study reported here aimed to address two questions: i) what are the perceptions and experiences of PwLC on accessing rehabilitation for Long COVID? and ii) what are GPs’ perceptions and experiences of managing PwLC presenting with symptoms of Long COVID that may be suitable for rehabilitation? PwLC were defined as adults (aged 18+) with Long COVID (with or without a positive COVID test) living in the community (i.e., not hospitalised).

## METHODS

### Study Design

This was a qualitative descriptive study employing semi-structured virtual interviews with a convenience sample of PwLC and GPs in four Scottish health boards. The study followed an a priori protocol (Supplementary File 1) and is reported in keeping with the consolidated criteria for reporting qualitative research (COREQ) [Supplementary File 2] [23].

### Participants

The four Scottish health boards were chosen for the larger realist evaluation study based on variation in population and accessibility (using the Scottish Government Urban Rural Classification 2020) [24], Long COVID prevalence, and Long COVID rehabilitation service delivery models. We recruited from these same health boards for this study. At the time of conducting this study, two health boards were offering an integrated Long COVID rehabilitation service (i.e., integrated within existing community rehabilitation pathways), one had recently launched a dedicated Long COVID community rehabilitation service, and one pre-existing dedicated Long COVID service was closed to new referrals due to an increased referral rate combined with reduction in funding and therefore inability to staff the service.

A convenience sample of PwLC were recruited via social media accounts of the research team and their institutions and by Long COVID Scotland, a volunteer-led charity run by PwLC. Inclusion criteria for PwLC were the following: community-dwelling (i.e., not currently hospitalised); aged 18 or over; experiencing symptoms of Long COVID (with or without a positive COVID-19 test; and experience of accessing or attempting to access healthcare services for possible rehabilitation. Those interested in the study contacted the research team, were sent detailed study information, and provided informed consent (audio recorded) prior to taking part. A second convenience sample of GPs were recruited by email invitation circulated on behalf of the research team by the NHS Research Scotland Primary Care Network. The network coordinator provided eligible GP practices in the four health boards with an electronic letter of invitation and participant information sheet to be shared with GPs. Those interested in taking part contacted the research team and provided informed consent as described above. Inclusion criteria for GPs were the following: GP in a practice within one of the four health boards taking part in the study; and experience of patients with probable Long COVID who may be suitable for rehabilitation. Several recruitment reminders were issued, and we aimed to recruit 12-20 PwLC and 8-20 GPs in total.

### Data collection

Interview topic guides [Supplementary File 3] were developed. While not formally pilot tested, the topic guides were refined by the research team, in consultation with people with lived experience of Long COVID and informed by initial findings from our realist evaluation study [15] and the wider literature in the field. During the period June 2022 to January 2023 semi-structured online interviews (Microsoft Teams) and one telephone interview were conducted by one research assistant [EHW], who received training and supervision from KC and ED. No repeat interviews were undertaken. GP participants were either in their workplace or homes when interviewed, while PwLC participants were all at home while interviewed. The research assistant, a female, was a qualified nurse and had worked in critical care from the start of the coronavirus pandemic (March 2020) until December 2021. She had no prior relationship with the GP practices or participants who all understood that EHW was employed as a study research assistant. The interviews lasted 17 to 47 minutes (mean 23 ±7.8 minutes) and were audio recorded. No other individuals were present during the interviews. No field notes for analysis were taken. Interviews with PwLC were transcribed by an external transcription service. Due to the homogeneity of the GP interviews, these were not transcribed but instead were listened to on multiple occasions by two researchers [25 26]. Neither transcripts nor audio files were returned to participants for comment or correction.

### Data analysis

Data were analysed using the framework method (Gale et al. 2013) [27], which proposes a matrix-based format to facilitate the sharing and management of data as a team. This approach is widely used in applied health research and recognised as appropriate for multidisciplinary research teams as it provides a structure to consider data within and across interviews. Familiarisation with the data began by reading and re-reading the transcripts (PwLC) and listening/re-listening to the recordings (GPs), making analytical notes that informed the ‘working analytical framework’ for each participant group [27]. Although line-by-line coding is common in qualitative research, it is also possible to develop a framework without engaging in explicit coding [28]; due to the small scale and nature of the data we adopted the latter approach. The framework method was used to construct matrices in Microsoft Excel, enabling the data to initially be summarised into broad categories. This was led by EHW, in close consultation with two experienced qualitative researchers (KC, ED). The charted data were then analysed by interpreting within and between participants to identify concepts, which were subsequently grouped into themes, and finally overarching themes consisting of data from both participant groups. This was an iterative process involving multiple researchers (KC, ED, JC, TT, JS, ES), the study team, and subsequent refinement until there was consensus that the data had been comprehensively analysed. As with our previous study we did not seek participant feedback directly on the findings. We did, however, present the findings in a webinar attended by health professionals and PwLC. Webinar attendees endorsed this study’s findings and reflected that the analysis reflected their personal experiences.

### Patient and public involvement

Two members of the public with lived experience of Long COVID were core members of the study team (AL, JO). Both contributed throughout the study and were integral to its success, helping with design and identification of important issues to explore. They co-developed study materials and interview topic guides and contributed to analysis and interpretation of study findings.

## RESULTS

### Participant characteristics

Twenty-five PwLC expressed an interest in taking part in the study. Of these 25, one did not meet the inclusion criteria for this study, but they were invited to take part in the larger realist evaluation. Thirteen PwLC did not pursue interest in participating in the study. Sixteen GPs expressed an interest, of whom three did not proceed to interview due to logistical challenges. Therefore, eleven PwLC (1 male, 10 female; aged 40-65 (mean 53)) and 13 GPs (5 male, 8 female) provided informed consent and took part in an interview. Figure 1 displays the numbers of participants recruited from each of the four health boards and Table 1 describes participants’ demographics. No participants who consented withdrew from the study.

**Figure 1:**
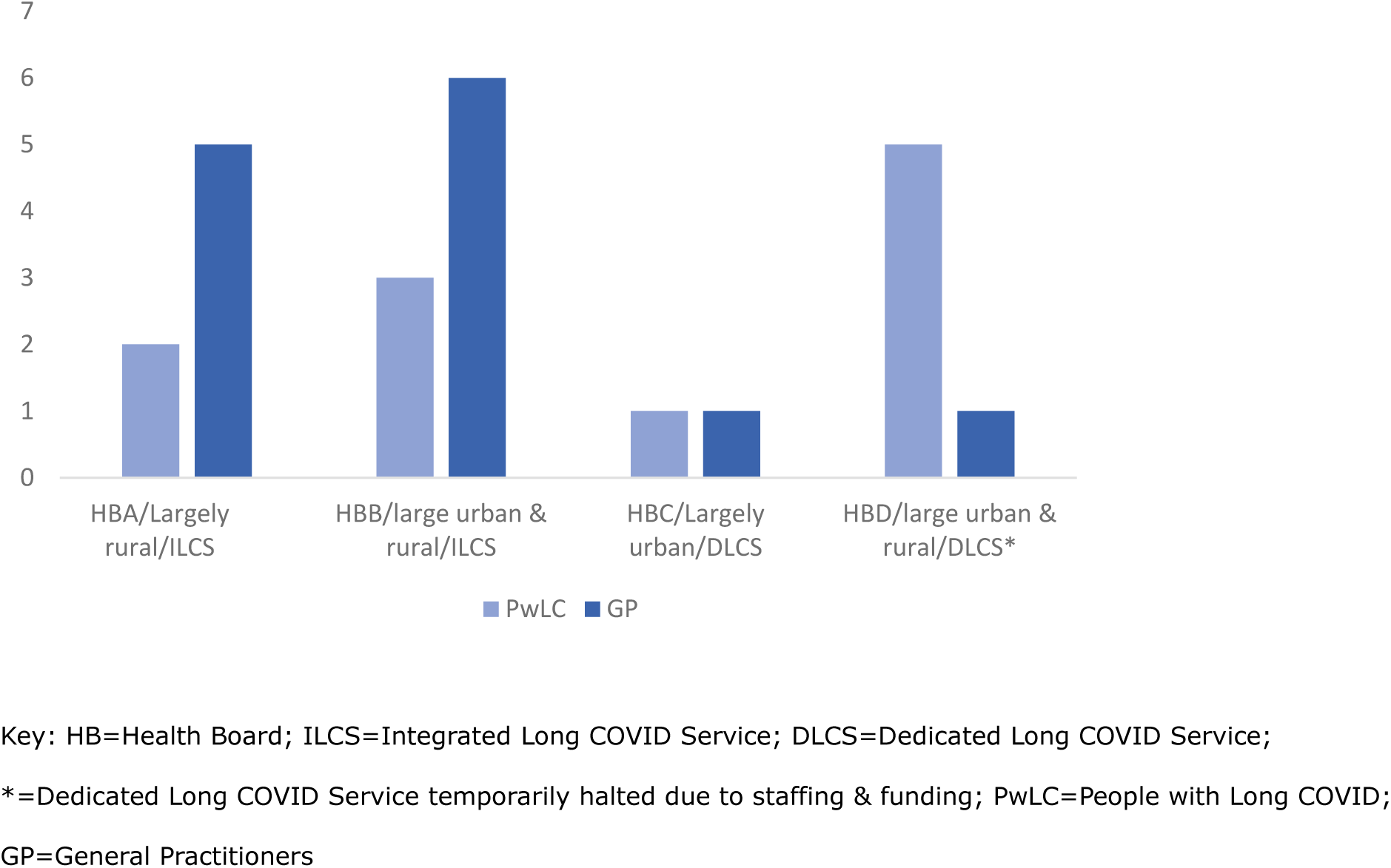
Number of GPs and People with Long COVID recruited from each health board.

**Table 1.**
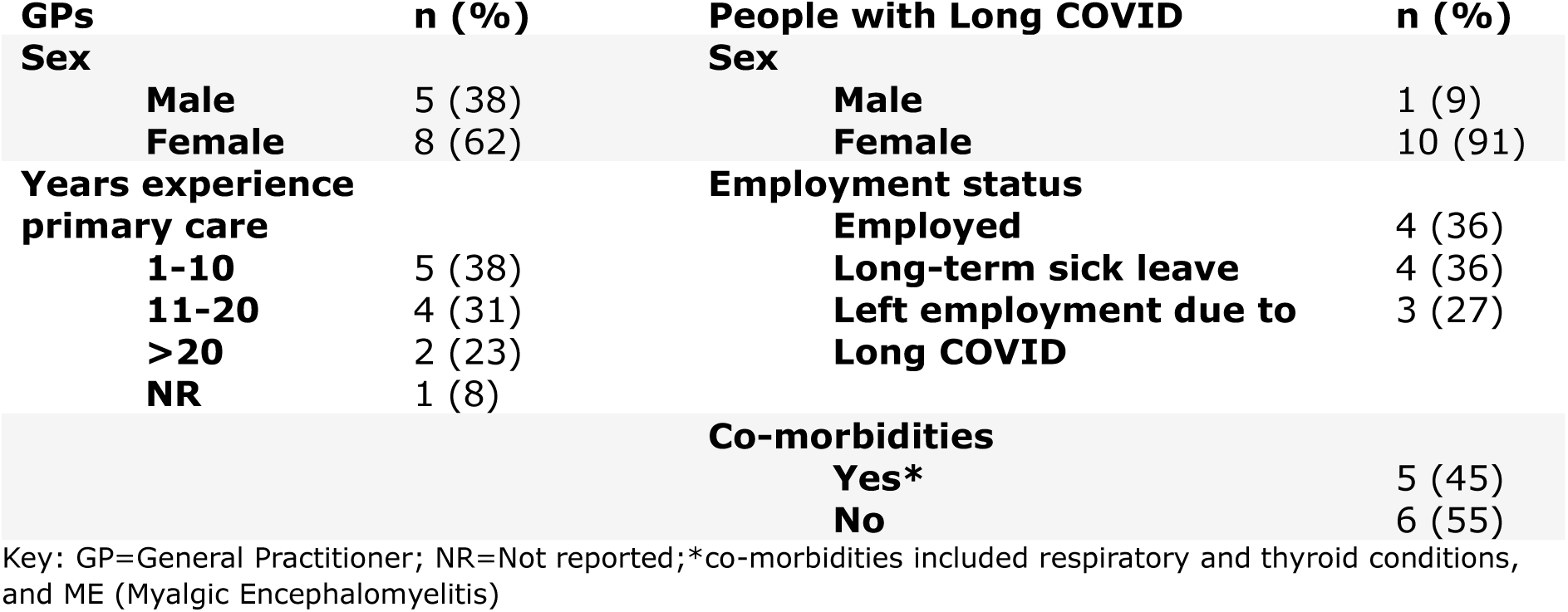
Participant demographics.

### Findings

Framework analysis resulted in four overarching themes: i) The lived experience of Long COVID; ii) The challenges of an emergent and complex chronic condition; iii) Systemic challenges for Long COVID service delivery, and iv) Perceptions and experiences of Long COVID and its management, including rehabilitation.

Table 2 identifies the data and participant groups that contributed to each of these themes. Throughout the manuscript participants are referred to by a unique, anonymous identifier (health board A-D, followed by PwLC or GP, followed by participant number).

**Table 2:**
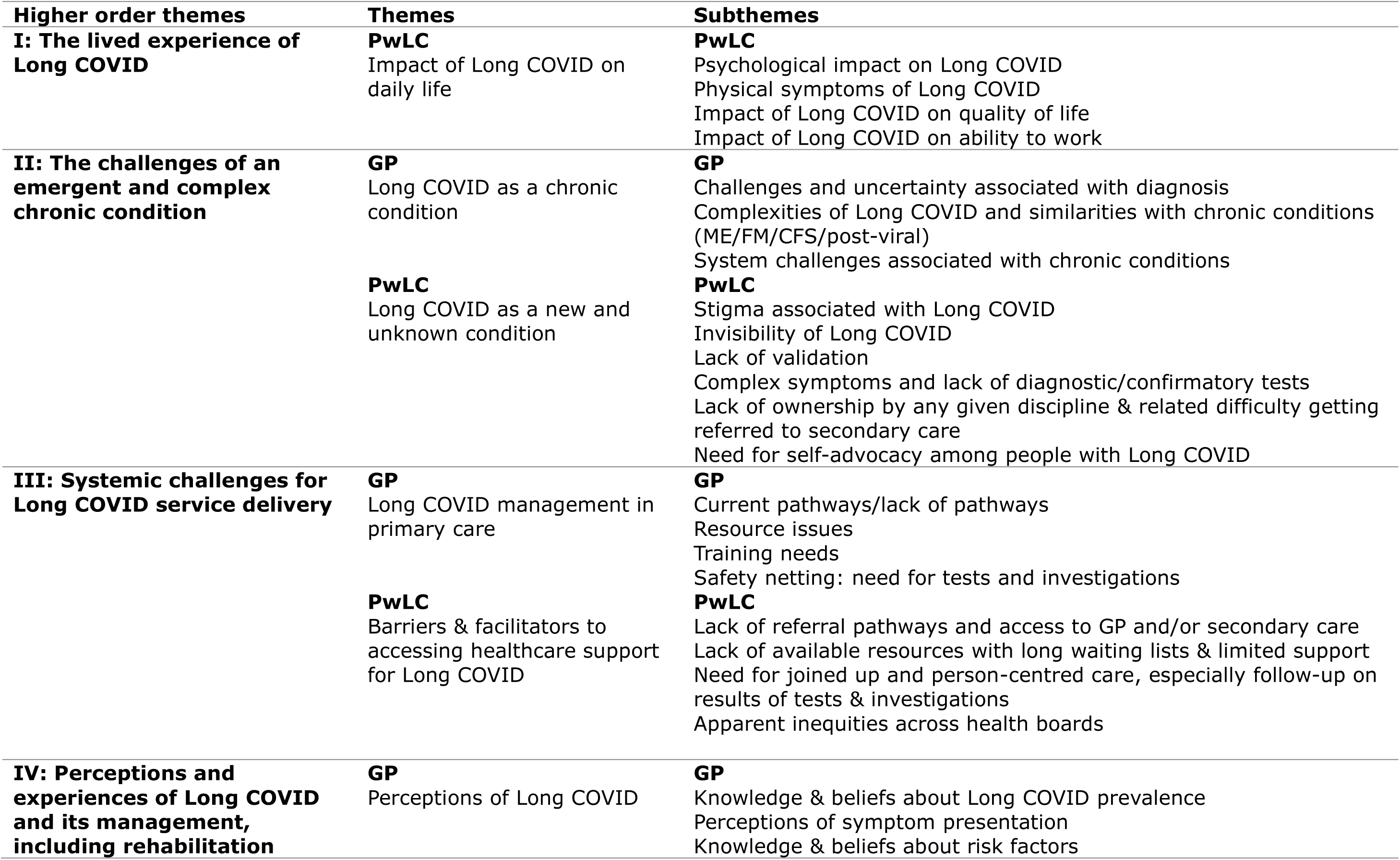
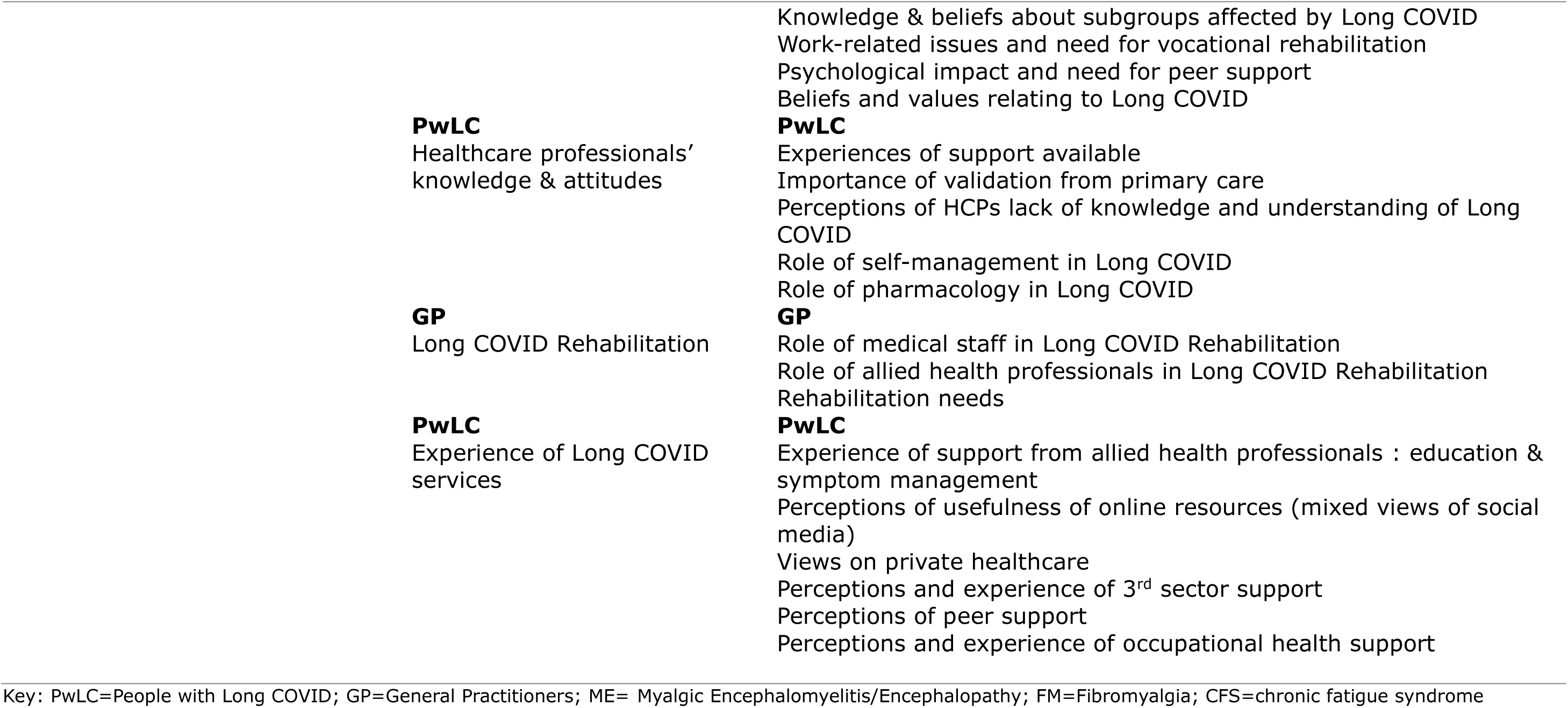
Overview of thematic analysis.

| **Higher order themes** | **Themes** | **Subthemes** |
| --- | --- | --- |
| **I: The lived experience of Long COVID** | **PwLC**<br>Impact of Long COVID on daily life | **PwLC**<br>Psychological impact on Long COVID<br>Physical symptoms of Long COVID<br>Impact of Long COVID on quality of life<br>Impact of Long COVID on ability to work |
| **II: The challenges of an emergent and complex chronic condition** | **GP**<br>Long COVID as a chronic condition<br>**PwLC**<br>Long COVID as a new and unknown condition | **GP**<br>Challenges and uncertainty associated with diagnosis<br>Complexities of Long COVID and similarities with chronic conditions (ME/FM/CFS/post-viral)<br>System challenges associated with chronic conditions<br>**PwLC**<br>Stigma associated with Long COVID<br>Invisibility of Long COVID<br>Lack of validation<br>Complex symptoms and lack of diagnostic/confirmatory tests<br>Lack of ownership by any given discipline & related difficulty getting referred to secondary care<br>Need for self-advocacy among people with Long COVID |
| **III: Systemic challenges for Long COVID service delivery** | **GP**<br>Long COVID management in primary care<br>**PwLC**<br>Barriers & facilitators to accessing healthcare support for Long COVID | **GP**<br>Current pathways/lack of pathways<br>Resource issues<br>Training needs<br>Safety netting: need for tests and investigations<br>**PwLC**<br>Lack of referral pathways and access to GP and/or secondary care<br>Lack of available resources with long waiting lists & limited support<br>Need for joined up and person-centred care, especially follow-up on results of tests & investigations<br>Apparent inequities across health boards |
| **IV: Perceptions and experiences of Long COVID and its management, including rehabilitation** | **GP**<br>Perceptions of Long COVID | **GP**<br>Knowledge & beliefs about Long COVID prevalence<br>Perceptions of symptom presentation<br>Knowledge & beliefs about risk factors |

|  |  |
| --- | --- |
|  | Knowledge & beliefs about subgroups affected by Long COVID<br>Work-related issues and need for vocational rehabilitation<br>Psychological impact and need for peer support<br>Beliefs and values relating to Long COVID |
| **PwLC**<br>Healthcare professionals' knowledge & attitudes | **PwLC**<br>Experiences of support available<br>Importance of validation from primary care<br>Perceptions of HCPs lack of knowledge and understanding of Long COVID<br>Role of self-management in Long COVID<br>Role of pharmacology in Long COVID |
| **GP**<br>Long COVID Rehabilitation | **GP**<br>Role of medical staff in Long COVID Rehabilitation<br>Role of allied health professionals in Long COVID Rehabilitation<br>Rehabilitation needs |
| **PwLC**<br>Experience of Long COVID services | **PwLC**<br>Experience of support from allied health professionals : education & symptom management<br>Perceptions of usefulness of online resources (mixed views of social media)<br>Views on private healthcare<br>Perceptions and experience of 3<sup>rd</sup> sector support<br>Perceptions of peer support<br>Perceptions and experience of occupational health support |

Key: PwLC=People with Long COVID; GP=General Practitioners; ME= Myalgic Encephalomyelitis/Encephalopathy; FM=Fibromyalgia; CFS=chronic fatigue syndrome

#### I: The lived experience of Long COVID

PwLC spoke of how their symptoms negatively influenced their quality of life. These included physical symptoms such as breathlessness and mobility issues, as well as cognitive difficulties such as ‘brain fog’ and problems concentrating. Fatigue was most frequently reported, with PwLC describing themselves as feeling physically and emotionally exhausted, resulting in a reduced capacity to participate in activities. Some PwLC were unable to engage in relationships or felt the need to reduce their level of engagement to preserve energy.

> *I was desperate, I just had no quality of life at all. I couldn’t speak to my friends for coughing, couldn’t look after my family because I had no energy, couldn’t get out the house, I was housebound for months. It was just rubbish.* [APwLC05]

Living with Long COVID had a profound impact on participants, with many describing the experience as significantly life changing. PwLC reported the deterioration in their physical health as leading to a sense of frailty.

This often caused frustration around their inability to participate in activities that were perceived as important, and a need to adapt and find new ways of managing daily life.

> *It is causing me to think now every time we decide to do something, ‘Am I going to be able? What are we going to have to do in order to facilitate that?’ And I’m not eighty-five. I wasn’t ready to have to start thinking about this right now.* [DPwLC05]

The psychological impact was characterised by PwLC as a struggle to come to terms with a new way of being, and a recognition that the internal expectations that one holds for oneself were being challenged.

This was described as a sense of losing or reshaping of one’s identity, with many acknowledging that they no longer felt like the person they used to be.

> *I can’t remember the last time when I had a day where I was thinking I feel like my old self…I think that last bit is the longest lasting effect of it and kind of the most debilitating, just feeling like a different and frankly lesser person.* [DPwLC06]

PwLC associated their physical health deterioration with a decline in mental health, with some PwLC experiencing negative feelings such as anxiety and depression. Participants also expressed a need to seek pharmacological and psychological support for their emotional wellbeing.

> *I also had - although it is something I suffer from, but I take Citalopram for anxiety- a very bad anxiety period as a result of COVID because to be honest I thought I was going to die, so that triggered a lot of other problems* [DPwLC05]

Living with Long COVID impacted some participants’ ability to engage in employment. Many attempted to return to work following the acute infection, with some returning to employment on a phased return. There was a perceived lack of support for return to work however, with some expressing a need for vocational rehabilitation to support return to work after a prolonged absence. Others spoke of the challenges associated with presenting at work whilst not able to fully function, which they attributed to a worsening of their physical symptoms.

> *Fortunately, I’ve had it, been off, but then went back to work. I’ve not stayed off. Maybe that’s a problem as well, I’ve not stayed off long term and listened to my body. I’ve went back and really threw myself back into my workplace and then suffered in my days off.* [CPwLC01]

The inability to work due to Long COVID created feelings of anxiety associated with the financial implications of losing income, and participants reported a lack of support and practical resources that could address their financial anxiety.

> *It is a worry for the future. I’m [age] and I’ve got a bit to go until retirement. I feel like I’m in limbo, it’s like Groundhog Day, and it’s kind of scary what’s going to happen with my work. I’m going to have to address that at some point because I can’t see there there’s an end in sight with everything. Yeah, it’s another hurdle I’m going to have to address next year at some point.* [BPwLC07]

#### II: The challenges of an emergent and complex chronic condition

Due to the symptoms experienced by PwLC in this study, many had presented to primary care seeking confirmatory tests and onward referral and had felt frustrated when they encountered GPs who were reluctant to provide a diagnosis. GPs however spoke of the challenges associated with providing a diagnosis in the absence of a specific diagnostic test, and the limited treatment options that were available.

> *I don’t know if we will move away from handing out the diagnosis of Long COVID.*

> *Because to be honest, I don’t often suggest it to the patient as a diagnosis because our options for management are so minimal. So, I tend to, if the patient thinks they’ve got it, work through that with them.* [AGP01]

GPs also drew similarities between Long COVID and existing chronic conditions, such as chronic fatigue syndrome/ME and Fibromyalgia, and a sense of familiarity around the uncertainties associated with diagnosis and management of these conditions, conveying a need for a service for long term conditions inclusive of Long COVID.

> *I think we are talking about Long COVID now because everyone is looking at COVID, which is great. But I think it’s not the only kind of post-[*illness*] treatment type problem that we have nowhere to send people. And I think it’s a problem with the health system.* [BGP01]

People with Long COVID recognised the uncertainties associated with Long COVID. Many felt they were not heard or believed by healthcare professionals and the wider public however and expressed a desire for their symptoms to be validated, listened to, and supported. There was a sense of social stigma associated with living with an invisible illness, which was often attributed to a lack of understanding about how Long COVID affects a person, and a feeling that they were being treated like a malingerer, and ‘making up their suffering.’

> *There’s definitely been a lot more media which is very helpful…there was a middle part where people were quite scathing because in the first part I remember friends saying ‘I don’t know anybody else that’s got Long COVID’, and that in itself felt a judgement, but then we hit a middle part where loads of folk were getting COVID but they’d been vaccinated or their bodies just dealt with it differently and they weren’t ill, so then there was a huge period of judgement came out, ‘well, so-and-so had it and they’re fine’.* [BPwLC06]

Many PwLC felt dissatisfied that symptoms were not being taken seriously enough to gain access to secondary care and had an awareness of inconsistencies across health boards in relation to Long COVID services, suggesting a need for a single person to take ownership of Long COVID in their area.

> *“There needs to be at least one thing for each trust. Even if there’s only one place in the whole of* [Health Board]*, and obviously I know that I’m from* [place] *and that would mean it would be here, I just think there should be at least one that people aren’t having to travel that long a distance to get to. Just somebody that has studied Long COVID, understands the complexities of it…* [BPwLC05]

The disconnection between patients’ expectations and their experiences led some to the realisation that they needed to take control of their own health. One participant stated, *“You’re almost left to fend for yourself”* [BPwLC07], whilst others highlighted the importance of advocating for themselves through researching symptoms, obtaining information from online resources, and using exercise to self-manage.

> *Oh, I’ve turned to Twitter…there is kind of a few people on Twitter that I follow that have been really good and kind of published research papers…I go back to the GP and ask about stuff, so I’m just having to kind of search for it myself…So you’re not only having to deal with the illness you have to kind of then navigate kind of like, where am I going to get help from?* [DPwLC02]

#### III: Systemic challenges for Long COVID service delivery

Healthcare system-related challenges included the lack of clear pathways to access, siloed specialised services, limited resources (long waiting times for appointments in primary and secondary care), and the lack of holistic care. PwLC described these challenges as contributing to communication breakdowns, a lack of accountability over chronic conditions, and the need for self-advocacy amongst people with Long COVID.

> *Every NHS trust should have at least one Long Covid clinic. It needs to have specialist people. All the therapists and all the scans and everything I’ve had have been done as standalone things, whereas at least if it was something centralised in a COVID clinic you would feel like you were having every part of your body tested and checked with Long COVID in mind....... It’s the piecemeal part of it that’s not looking at it in a holistic way.* [BPwLC05]

GPs reported a lack of pathways and processes for managing patients with Long COVID, including identifying and tracking patients along the care trajectory and establishing consistent approaches to management. The nature of the emergent condition made it challenging to identify appropriate services to refer patients on to.

> *It’s less easy to identify one individual you could refer them to so without an over-arching service it’s hard to know where to direct the patients.* [AGP02]

GPs reported that patients seldom met the criteria for existing rehabilitation programmes (e.g., cardiac/respiratory). They also reported that they often carried out tests and investigations to exclude other differential diagnoses as a means of ‘safety-netting’, but some were mindful that this could delay rehabilitation and recovery.

> *There is a risk that we hold up rehabilitative inputs until we fully investigated things and we are entirely assured that there’s nothing going on. So, we should perhaps be blending things a little better.* [BGP02]

Both participant groups felt that limited resource was one of the main barriers to Long COVID management. People with Long COVID experienced difficulty accessing both primary and secondary care.

> *I didn’t have the energy to argue with the receptionist at the GP surgery, and that’s the honest truth, I just didn’t have it in me to phone and try and explain it all again. So, I lost nineteen pounds in four weeks because I just couldn’t eat because I felt sick. But even that wasn’t worth the battle I would have to get past a GP receptionist.* [APwLC05]

GPs were aware of the significant pressure secondary care were facing which often resulted in long waiting times for specialties. As a result, patients often re-presented to primary care for ongoing issues leaving GPs with limited options for referrals and treatments.

> *We were getting a bit frustrated referring patients to secondary care for help. There wasn’t much coming through. They were already dealing with backlog enough and they’re getting piled up with these other things happening. I understand their limitations, entirely…I don’t think that any I have referred* [to respiratory or cardiovascular] *have been seen yet. …* [CGP03]

Variation in models of access for rehabilitation led PwLC to question the equity of care across health boards as they were aware of Long COVID services provided in other parts of Scotland and in England. Some PwLC who were able to afford it sought private health care and experienced improvements in their condition.

> *I’ve taken it upon myself to see a physio privately who is helping me because that’s the hardest thing is this pacing business…Because I do too much one day, then I can’t do anything the following day, maybe even two days…It’s been very helpful because she, she’s guiding me because what I would I think is manageable and doable is very different to what she says is manageable and doable.* [DPwLC01]

People with Long COVID emphasised the need for a more joined-up service to improve communication and coordination of care.

> *I should have had the results in six weeks, but I’ve not had them, I don’t know who to contact to get them, because it was through the neurologist rather than my GP.* [APwLC03]

They also felt that specialists focused only on specific aspects of their health but not the condition as a whole.

> *I just feel sometimes having that one person, like I know a lot of people have a consultant that they go to and that’s the person that they speak to, or the centre that they go to, for support. There’s a group of people perhaps that they deal with, but they get it. They’re a familiar face and that makes sense. But you feel a bit of a pariah, to be honest, with Long COVID.* [BPwLC06]

GPs acknowledged the importance of a more holistic approach to Long COVID care including psychological as well as physical support. They suggested the need for integrated multidisciplinary management that provides support for a complex range of symptoms.

#### IV: Perceptions and experiences of Long COVID and its management, including rehabilitation

This theme comprised data on GPs knowledge, attitudes, and perceptions of Long COVID, as well as participants’ perceptions and experiences of Long COVID services.

##### GPs knowledge, attitudes, and perceptions of Long COVID

In health boards where Long COVID management was integrated into existing community services with less clear pathways to rehabilitation (health boards A & B), there was a perceived lack of demand by GPs for Long COVID-specific services. GPs in this study reported low numbers (1-6 per week) of people presenting to their practices for support with symptoms of Long COVID.

> *Maybe 10-12* [Long COVID patients have presented] *in total. But I don’t know whether they’re all coming to us. They might be just suffering in silence.* [CGP03]

Patients were commonly reported to be of working age and were perceived to be fit and healthy prior to coronavirus infection. Some GPs also reported that females were more commonly presenting with symptoms associated with Long COVID than males.

Some GPs attributed the low patient numbers to natural resolution and the ability to self-manage on the basis that patients were not reconsulting or requesting further sick lines.

> *I think with time most of their symptoms do seem to ease and pass.* [AGP01]

However, some GPs and most PwLC believed that people with Long COVID were not presenting to primary care because they believed there was no support available for Long COVID.

> *A lot of patients don’t necessarily consult because probably they are seeing things in the media and things, you know aware that there aren’t particular treatments. So, they just think it’s par for the course that they feel like that.* [AGP02]

> *“I stopped contacting the GP because I just feel I’m wasting their time”*

> [APwLC03]

PwLC expressed some concerns associated with GPs being under pressure and attributed their reduced attendance at GP practices to feeling like they were an additional burden on the healthcare system. They spoke of withholding information related to their symptoms due to an awareness that GPs were time pressured, and expressed concerns associated with ensuring appointments were productive.

> *I know there are things that I haven’t raised with a GP because I’m aware that they’re time pressured, I’ve raised about ten symptoms already in my consultation with them and I know I’ve got another three sitting on my list, but I can’t bring that into the situation…and I’m potentially sitting on stuff that I should have discussed* [BPwLC06]

PwLC also noted their concerns over the lack of specialist knowledge and expressed a sense of apprehension when GPs were unable to demonstrate an appropriate level of knowledge concerning the condition.

> *But the last time I went to the GP they said, ‘we find people with Long COVID know more about it than we do.’ And I thought that doesn’t really fill you with great enthusiasm.* [DPwLC04]

Most GPs acknowledged that Long COVID has impacted people living with the condition and conveyed a need for education associated with recovery and returning to work. They spoke of patients feeling pressured to return to work without appropriate support in place to assist their workplace integration. The need for psychological support was also expressed by both GPs and PwLC. Specifically, peer support was identified as a useful resource for people with Long COVID, where they can be supported by others experiencing similar symptoms.

> *It’s just reassuring to know that you’re not alone in this. Misery loves company, and it’s good to know that there are other people who have this, because otherwise it would become kind of depressing. And it helps put things in perspective, that you know that as bad as you feel someone else is probably feeling worse.* [DPwLC06]

A Minority of GPs did not perceive the need for a specialised Long COVID service. Some GPs referred to a scepticism among their colleagues about Long COVID in general:

> *It’s not only for people, it’s also for the medical professionals to believe as well that this is a problem. I think there is still some scepticism among medical professionals as well, still, about this being accepted and treated.* [CGP03]

One GP participant described themselves as being slightly cynical about Long COVID and wary of the need for rehabilitation, suggesting that patients should *“wait it out”* and they will get better, likening Long COVID and its impact to ‘flu.

> *“I think it’s exactly like flu. The same applies in flu. You get lots of people that get it. Most people are not terribly well with it, few people get flu without knowing they’ve had it. Some people recover quickly, some take a longer time to recover and some die”* [AGP04]

##### Perceptions and experiences of rehabilitation and other Long COVID services

A range of perceptions regarding Long COVID rehabilitation were held by both participant groups. Some PwLC lacked knowledge of the potential role of rehabilitation professionals in supporting people with Long COVID. This view was shared by some GPs who reported a limited understanding of the role of rehabilitation in the management of Long COVID.

> *Physio, I can’t really see much of a role. But that could be my lack of knowledge about it because with the patients I have spoken to it’s not really so much of a physical thing, it’s not like a particular joint pain as such that they would benefit from a physio. It’s more the kind of cognitive aspect, maybe an OT, but I don’t know what they would add.* [BGP05]

Some PwLC had enough knowledge of rehabilitation to request referral or refer themselves to rehabilitation services where this was an option. In most cases these participants were on waiting lists and had been for some time. Although we did not record participants’ job titles (for those in employment), several participants in this category disclosed during the interviews that they were healthcare professionals, with prior knowledge of, or colleagues working in, rehabilitation services. A third group of PwLC reported a need for Long COVID rehabilitation services in their health board area, suggesting a lack of services and/or their promotion.

The small number of participants who had received rehabilitation for Long COVID (n=3) were generally positive about their experience, reporting benefits from specific professions (e.g., physiotherapy, speech and language therapy) and interventions (e.g., breathing exercises), with information and advice on Long COVID and symptom management, particularly the use of pacing, being highly valued.

> *The biggest help was speaking to Speech and Language… she gave me lots of information that was very interesting, and lots about the biology of what’s going on with my* [laryngeal] *spasms.* [DPwLC04]

> *Just having somebody to help you manage what that should look like, what is too much, because you can read about pacing, you can chat about it online with other people with Long COVID, but trying to get a model that fits for you as an individual is actually really hard without support.* [DPwLC06]

This contrasted with the views of PwLC of generic self-management booklets, which were commonly reported as lacking person-centredness.

> *When I did refer myself to the* [specialist] *team, I got a booklet, a massive booklet through the post, that says this that and the other. But it’s such an individual, highly differentiated set of symptoms that any one person can have, just none of it was particularly relevant to me.* [DPwLC01]

One participant had sought informal rehabilitation from a personal trainer but felt that was not helpful as it was too intense, and the trainer did not have the requisite Long COVID knowledge to support patients effectively to avoid rehabilitation being ‘too intense.’

> *“Just with having this discussion it’s like a wee light bulb moment that I’m having, that I’m thinking I’ve tried the PT* [personal training]*, it was too intense, threw the towel in. See all that weights and lunges and dah, dah, dah, I thought I haven’t got the energy for it. So maybe physio, as you’re saying, if they know all about the disease, they know about what they would recommend and maybe be a bit more gentle”* [DPwLC01]

Despite the lack of knowledge of the potential role of rehabilitation demonstrated by some GPs, most reported that they would engage with a dedicated Long COVID rehabilitation service, as it would provide an onward referral route for patients, particularly as they commonly reported being limited by secondary care referral criteria. Several GPs felt that a multidisciplinary team approach could be beneficial for their patients and could provide the validation that patients needed.

> *I think they feel quite isolated actually and I think it would be useful even if objectively…there’s not a huge improvement. I think psychologically it would be really important for them. Someone to believe them, to see what’s happening, and just thinking someone’s looking out for them.* [BGP01]

Some also believed that earlier pulmonary rehabilitation could contribute to better functional recovery.

PwLC reported accessing a range of other Long COVID services and sources of support. For some, a helpful source of advice and support came from occupational health services when they were returning to work after a period of Long COVID-related absence. Some PwLC had accessed support from a charity, with some finding it useful and others finding it too generic for their needs. Some participants found support from peers with similar experience of Long COVID was helpful and provided validation. For some this positive peer support was accessed via social media. Others reported negative experiences of social media including finding social media platforms unsafe and unhelpful for sharing lived experiences. The lack of monitoring in online forums contributed to PwLC feeling vulnerable to receiving incorrect information and negativity from others. Many PwLC reported that they resorted to online information, with some taking this information to their GP consultation. Finally, as reported above, some turned to private healthcare in response to access issues and long waiting times, including psychological therapy, GP, medical specialties, and physiotherapy.

## DISCUSSION

We explored the issue of access to community rehabilitation for Long COVID from the perspectives of PwLC and GPs in four Scottish health boards. PwLC described a range of symptoms affecting their physical and mental health and having a profoundly negative impact on their daily life, employment, and relationships. We found a tension between PwLC seeking diagnosis and onward referral and GPs lacking diagnostic tests and limited options for managing Long COVID. This tension could manifest itself in PwLC perceiving themselves as socially stigmatised by their ‘invisible’ condition. We also identified several systemic challenges for Long COVID service delivery which related to access, siloed services, limited resources, and a perceived lack of holistic care, causing frustration for both GPs and PwLC. We found that although a minority of GPs expressed scepticism about Long COVID and the need for rehabilitation and other services for this patient group, there was general agreement between PwLC and GPs on the need for accessible, person-centred services and support. Regarding community rehabilitation, we found that: i) some PwLC and some GPs lacked knowledge on the potential role of community rehabilitation in the management of Long COVID; ii) having prior knowledge of rehabilitation or being a healthcare professional appeared to facilitate access to community rehabilitation, and iii) PwLC who had received rehabilitation generally found it beneficial. Due to the lack of knowledge and difficulty accessing rehabilitation however, PwLC had accessed a range of other services and sources of support, with varying success.

The relationship between Long COVID and reduction in physical and mental health outcomes and quality of life is becoming increasingly known [29 30]. A large Scottish population cohort study found that almost 50% of PwLC had not recovered 6-18 months after symptomatic infection, and that PwLC experienced a wide range of symptoms affecting daily activities and quality of life [31]. This study adds to that growing evidence-base and demonstrates the influence of the condition on a sample of PwLC living in Scotland almost 3-years on from the start of the pandemic. The persisting prevalence and impact of Long COVID on people’s lives further emphasises the need for support and services such as community rehabilitation to be available, in keeping with recommendations [1 6 8].

These recommendations are that PwLC should have access to personalised and multidisciplinary rehabilitation; such rehabilitation is reportedly available throughout Scotland [12] delivered by a variety of service models. Our previous research, however, observed low numbers of PwLC receiving community rehabilitation in some areas of Scotland [15]. This study has shed some light on the potential reasons for the mismatch between recommendations, reported service availability and numbers of PwLC accessing community rehabilitation.

Lack of GP knowledge regarding community rehabilitation and its potential role in Long COVID may, in part, be attributed to the nature of Long COVID as a new condition that health professionals are still learning how to manage. Previous research has reported a lack of GP understanding of the role of rehabilitation professionals in the management of conditions commonly encountered in primary care; for example, the role of physiotherapists in osteoarthritis management [32]. Furthermore, international public understanding of the role of occupational therapy has been reported as limited [33]. Therefore, in the context of a new condition with an evolving evidence-base, it is perhaps not surprising that GPs and PwLC may have limited understanding of what rehabilitation professionals can offer to this patient population. Furthermore, the reluctance to promote the availability of services in some areas, due to pre-existing resourcing and anticipated demand-capacity issues may have further limited access to Long COVID rehabilitation [15]. Issues with promotion of services, clarity of pathways and interdisciplinary communication between GPs and rehabilitation professionals have been reported previously[32 34]. The findings of this study suggest that further improvements in communication and collaborative working may be required to enhance access to community rehabilitation for PwLC [32]. Indeed, the finding that prior knowledge of rehabilitation, and being a healthcare professional facilitated access to community rehabilitation for PwLC, is further evidence that successfully navigating the referral system is challenging.

The findings of PwLC who had managed to access community rehabilitation being satisfied with it, and GPs wanting a Long COVID service to refer patients to are in keeping with previous research [17]. The challenge not only lies in availability of such services, but clearly in PwLC and GPs awareness of the benefits of these services, their active promotion, and clear and timely accessibility of rehabilitation services. There is therefore a need to overcome the systemic challenges to accessing timely rehabilitation reported in this study. Considering the ongoing nature of living with Long COVID, these challenges are likely to continue beyond the time and resource constraints of Government funding provision for existing Long COVID rehabilitation within rehabilitation services that were already historically underfunded and considered ‘Cinderella services’ [15].

### Strengths and limitations

This was the first study to explore the issue of accessing community rehabilitation in the Scottish context and included the perspectives of those referring and potentially being referred to community rehabilitation. One researcher conducted all interviews to ensure consistency and multiple researchers were involved in analysing and interpreting the data, including people with lived experience of Long COVID. There are, however, some limitations. As this study was an extension of our original study, we had insufficient resources to enable larger participant samples. We realised from the outset that we would be unable to describe achieving data saturation. Although we recruited to target for the GP sample, there was under-representation from the two health boards with dedicated Long COVID services (one newly launched and one halted). Therefore, the data largely represents the views of GPs from health boards where Long COVID services were integrated into existing community rehabilitation services; it is possible that GPs views of dedicated Long COVID services may be different. We recruited a small, mostly female sample of PwLC, and mostly from health boards with integrated or a halted dedicated Long COVID service. Their views on accessing Long COVID services may therefore have been biased. Recruitment of PwLC was likely limited by our reliance on social media and one Long COVID charity; however, both mechanisms had the potential to reach many people. Despite these challenges, both the PwLC and GP participant groups provided consistent understandings of the challenges they faced when dealing with Long COVID. Therefore, while data saturation is not claimed, it does appear that we achieved adequate data sufficiency to be confident that the findings reflect key issues within each participant group.

### Implications for practice and research

There is a need for greater understanding by the public, GPs, and other potential referrers of the role of community rehabilitation professionals in the management of Long COVID. There is an equally important need for community rehabilitation services to be well promoted and accessible to the PwLC for whom they may be appropriate. Long COVID is still a prevalent condition whose impact on individuals can be profound. The need for community rehabilitation for PwLC is likely to persist. Service providers should therefore consider availability and accessibility of Long COVID rehabilitation and ensure adequate interprofessional communication and collaboration to enhance the experience for PwLC.

## CONCLUSION

We have provided further understanding of the barriers to accessing Long COVID community rehabilitation by exploring the perceptions and experiences of key stakeholders in the referral process. These findings can be used by those (re)designing community rehabilitation services for people with Long COVID and potentially for other conditions. There remains a need for greater public and GP awareness of the role of rehabilitation professionals in Long COVID.

## AUTHOR CONTRIBUTIONS

KC, ED, JM, LA, JP, JO, AL, PS contributed to the study’s conception and design. EHW undertook data collection. ED, KC, EHW, TT, JC, JS, ES undertook analyses and drafted the first version of the manuscript. KC led future iterations of the manuscript. All authors read and commented on the manuscript and approved the final version of it. The corresponding author attests that all authors meet authorship criteria and that nobody meeting the criteria have been omitted. ED and KC have joint responsibility for the research conduct of the study, had access to the data, and controlled the decision to publish.

## DATA AVAILABILITY

All data produced in the present study are available upon reasonable request. Robert Gordon University holds the copyright for the full interview transcripts and may grant data sharing permission on request.

## ETHICS APPROVAL

Ethical approval was granted from the Wales Research Ethics Committee 6 [21/WA/0118 A03], and each of the four Health Boards granted R&D management approval.

## ACKNOWLEDGEMENTS

We are grateful to all our participants who gave of their time to take part in interviews. We would like to thank Amanda Cardy, Northeast coordinator NRS Primary Care Network and local principal investigators Lynne Frew, Lynn Morrison, and Gail Thomson-Patel for support with participant recruitment.

