## Supplementary File 1 for "Exploring the perceptions and experiences of community rehabilitation for Long COVID from the perspectives of Scottish General Practitioners’ and people living with Long COVID: a qualitative study" (from another version of this preprint: version 1, 2023-11-06)

**IRAS ID: 295672**

**Protocol**

### 1.0 Title

Evaluating long-covid rehabilitation in Scotland

### 2.0 Summary

Studies estimate that 10–35% of people with COVID-19 experience disabling clinical symptoms following the acute phase of their illness (long-covid). Given the prevalence of COVID-19 in Scotland, there is assumed to be a significant number of people experiencing long-covid, and they are starting to be referred to rehabilitation specialists. Community rehabilitation, delivered by physiotherapists, occupational therapists and other health professionals, is a well-established approach to enabling people to maximise their quality of life and recovery. However, community rehabilitation for people with long-covid is in its infancy. There are considerable differences in how community rehabilitation for people with long-covid is currently delivered across Scotland. We want to assess which models of community rehabilitation are most appropriate, in which circumstances. We will evaluate the delivery and outcomes of four different models of community rehabilitation for people with long-covid currently being delivered in Scotland. This will help us to identify which models are more suitable, for which patient groups and in which contexts. We will then hold online workshops with community rehabilitation managers, service leads and others from across Scotland. During the workshops, we will present our findings and support participants to develop evidence-based action plans to improve their local long-covid community rehabilitation services

### 3.0 Background

#### *3.1 Introduction*

This study is an in-depth exploration of models being used to deliver rehabilitation for longer-term effects of COVID-19 in Scotland. Understanding and evaluating how service models are developing, in parallel with evolving knowledge of the impact of COVID-19 on patients, will inform the development of new and responsive patient-centred models that can effectively meet patient and service need. This work is vital. As evidence emerges on the epidemiology and consequences of COVID-19, its impact on individuals and society, and potential burden on health services (1), strategies are required to address the long-term effects of COVID-19. The Scottish Government's Framework for Supporting People through Recovery and Rehabilitation during and after the COVID-19 Pandemic (2) states that "everyone with rehabilitation needs…will be able to access the care and support they need to live well, on their own terms." It also highlights the important role of digital solutions. This research will develop implementable recommendations for post-COVID-19 rehabilitation, congruent with the principles of the Scottish Rehabilitation Framework and Digital Health and Care Strategy (3), and that meet patients' needs.

#### *3.2: Long-covid*

Long-term effects of COVID-19, or "long-covid" has received much recent media attention with action groups calling for more support (4). With no agreed definition, we will use that proposed by Greenhalgh et al (5) of symptoms extending 3-weeks beyond onset (post-acute COVID-19) and 12-weeks beyond onset (chronic COVID-19). Studies on long-term consequences of COVID-19 infection are ongoing; however, it is estimated that 10 – 35% of patients may experience post-acute COVID-19 (5-7). Based on Scottish Government figures for positive COVID-19 tests and survival to date (28th February 2021) this equates to between 19,367 and 67,784 people in Scotland with post-acute COVID-19, with around 2,000 remaining significantly unwell at 12-weeks, commonly due to organ damage (7). Long-covid also affects people with milder symptoms and those who were not hospitalised (6). Rehabilitation therefore needs to be accessible to those presenting in community settings as well as being discharged from hospital.

Reported symptoms vary widely and commonly include respiratory, cardiopulmonary, neurological, musculoskeletal and mental wellbeing sequelae, as well as fatigue and loss of taste and smell (5,6,8). As described above, presentation and severity are variable, and it is important to note that many patients with long-covid will have co-existing conditions that will impact on symptoms and require dual management. Rehabilitation therefore needs to be multidisciplinary, comprehensive, and tailored to patients' needs, in order to maximise function, quality of life and participation in society (9).

#### *3.3: Long-covid rehabilitation*

Rehabilitation for long-covid is in its infancy, reflected in a recent living systematic review update finding predominantly expert opinion, indicating that high-quality research is required (10). Research on rehabilitation for other respiratory conditions *may* be generalisable to the COVID-19 population (11), but this is yet to be determined. Several UK expert bodies and published expert opinion recommend a stepped, needs-based rehabilitation approach incorporating information provision, self-management support and specialist services as required. They also recommend that rehabilitation should be individualised and progressive and the use of digital solutions optimised. *How* rehabilitation can be optimally delivered is not known but vital to determine, given that rehabilitation services need to cope with additional COVID-19 demand whilst continuing to provide rehabilitation for other, often vulnerable, patient populations (12). By investigating different ***models of community rehabilitation*** currently implemented in Scotland to evaluate what works, where, with whom and how, we will develop recommendations on *how* rehabilitation can be best delivered across Scotland to meet the needs of patients and services, thereby developing new responsive models of rehabilitation.

### 4.0 Aims & research questions

The overarching aim of this research is to provide responsive evidence-based recommendations to the NHS about how to most effectively deliver community rehabilitation for people with long-covid. We will do this by evaluating the implementation and impact of different emerging models of long-covid rehabilitation in different contexts in Scotland. We will address the following specific research questions:

1. How is rehabilitation for long-covid being delivered in 4 Scottish health boards?
2. What are the barriers and facilitators to implementing long-covid rehabilitation from managerial, rehabilitation staff, and patients' perspectives and experiences?
3. What are the perceptions and experiences of community-dwelling adults with long-covid on accessing rehabilitation for long-covid?
4. What are GPs perceptions and experiences of managing community dwelling adults presenting with symptoms of long-covid that may be suitable for rehabilitation?
5. What are the outcomes of different community rehabilitation models for people experiencing long-covid?
6. What are the costs associated with long-covid rehabilitation in the 4 selected cases?
7. In what ways are the selected long-covid community rehabilitation models successful (or not)?
8. How can we use the knowledge from 1-5 above to inform the development of responsive patient-centred models of rehabilitation that can be adapted to suit the needs of patients and services across Scotland?

### 5.0 Methodology & methods

We will conduct a mixed-methods case study realist evaluation (RE) of different models of community rehabilitation, in different contextual settings, for people experiencing long-covid in order to understand what works, for whom, in what circumstances and how. The study is informed by the RE framework (13), which is used to study the process and context of 'implementation' and to identify the 'mechanisms of action' that lead to successful (or unsuccessful) delivery. It will also be used to investigate how interventions are being implemented in each case study area. Understanding the local contexts and mechanisms of action will enable us to explain observed outcomes in the different models of community rehabilitation delivery for people with long-covid. Realist Evaluation is a highly appropriate method to use to study the delivery and impact of emerging models of care for long-covid. Realist evaluation emerged in response to the need for knowledge that extends beyond that obtained by controlled trial design (is the intervention effective?) to answering questions of what works for whom and in what circumstances. It is 'realist' in the sense that it does not deny the relevance of robust measurement of outcomes, but also seeks to take account of the context in which interventions are delivered and to identify the mechanisms that make programmes work (or not) to produce the observed outcomes. It is not the interventions themselves that work but the ability and willingness of participants to engage with them.

***Setting***

Rehabilitation services in four Scottish Health Boards: NHS Ayrshire & Arran, NHS Grampian, NHS Lanarkshire, NHS Tayside. The boards have been selected to represent a diversity of models of rehabilitation in terms of mode of delivery (digital/face-to-face/blended/self-management) and whether they provide a new long-covid service or integrate long-covid rehabilitation into existing services. This information was gained by a Scottish-wide survey of long-covid rehabilitation conducted by the study team (14). The selected NHS boards also represent a range of geographical areas in relation to urban/rural classification, to allow for transferability of findings across Scotland, and boards with sufficient covid-19 cases to make recruitment viable.

The study consists of 2 work packages (WP):

**WP1:** Case study RE to measure implementation and outcomes of different service models of community rehabilitation for people experiencing long-covid. Within WP1 we will conduct:

***I. Process evaluation addressing research questions 1-4***

Data will be collected at 4 time-points over a 12-month period across each case to understand local contexts and decisions regarding delivery of community rehabilitation for people with long-covid. Data collection rounds 1 to 4 will describe the long-covid models of community rehabilitation and track local decisions on how services are implementing and developing their long-covid rehabilitation service. Data collected during rounds 1 to 4 will elicit rationales for service delivery decision making and how implementation is supposed to work. Data collection will be obtained from online focus groups (with people who have received long-covid community rehabilitation), and online individual interviews (6-10 per case per round) with managers/service leads/consultants/ and rehabilitation staff with potential to deliver long-covid community rehabilitation. Data from Round 1 will specifically be used to elicit rationales regarding local decisions, identify 'folk theories' surrounding how community rehabilitation delivery is supposed to work, and identify hypothesised mechanisms of (successful) change/implementation. This will establish the locally hypothesised context-mechanism-outcome (CMO) configurations which will then be tested using data gathered in Rounds 2-4.

***Participants***

1. People who have received long-covid rehabilitation (patients), managers/service leads, and professionals delivering long-covid rehabilitation will be recruited. See table 1 for inclusion/exclusion criteria.
2. Community dwelling adults with long-covid (i.e., not currently receiving community rehabilitation) and GPs in each of the four health boards.

***Recruitment***

***Patients*** will be sent a study pack in one of the following ways: (i) with their long-covid appointment if it is mailed out to them; in-person from their rehabilitation professional when they receive their first session (either in their home or in a community rehabilitation setting); by mail if their initial session is held virtually. The study pack will consist of: i) letter of invitation, ii) participant information sheet, iii) baseline measures. Patients will therefore be identified by administrative or clinical staff, and will opt-in to the study by completing the baseline measures (paper-based or online) and returning them to the research team. Participants will opt-in to taking part in a focus group, and if they wish to do so will provide contact details for the study team to contact them.

**Table 1: Inclusion/exclusion criteria participants**

| | **Inclusion** | **Exclusion** |
|---|---|---|
| **Patients** | -Aged 18+<br>-COVID-19 diagnosis (with or without positive covid-19 test) >3-weeks previously<br>-Referred due to presenting with symptoms thought to be due to long-covid (including but not limited to: respiratory, fatigue, musculoskeletal, mental health)<br>-Suitable for rehabilitation (determined by rehabilitation staff)<br>-Able to understand written & spoken English<br>-Willing to take part in study | -Aged <18<br>-Requiring specialist (inpatient) management of specific complications<br>-Unable or unwilling to provide informed consent<br>-Other life-limiting illness with life expectancy <6-months e.g. disseminated malignancy |
| **Managers/service leads** | Managers/service leads of community rehabilitation services providing long-covid rehabilitation | Managers/service leads of inpatient services |
| **Rehabilitation professionals** | Healthcare professionals delivering long-covid rehabilitation including but not limited to, physiotherapists, occupational therapists, speech and language therapists, psychologists, medical doctors. Where relevant (i.e. involved in delivering long-covid rehabilitation) healthcare support workers will be included | -Student healthcare professionals<br>-Healthcare professionals providing in-patient rehabilitation |
| **Community dwelling adults with long-covid** | Aged 18+<br>Self-identify as having experience of long-covid and accessing/attempting to access healthcare services for possible rehabilitation | -Aged <18<br>-Unable or unwilling to provide informed consent<br>-Other life-limiting illness with life expectancy <6-months e.g., disseminated malignancy |
| **GPs** | GP based in any of the four participating health boards with experience of patients with probable long-covid who | |

| | |
|---|---|
| | may be suitable for rehabilitation |

***Rehabilitation professionals and Managers/service leads*** will be recruited by email with a letter of invitation and participant information sheet attached. The email will come from a relevant gatekeeper e.g. team lead/manager for rehabilitation professionals; sector lead/allied health professions lead for service leads/managers. Participants will opt-in to the study by contacting the research team by email or telephone.

**Community dwelling adults with long-covid** will be recruited via social media using RGU School of health sciences and NMAHP research unit University of Stirling Facebook and Twitter accounts, via long-COVID Scotland action group, and The Alliance. Following these routes, we anticipate snowball sampling will occur.

**GPs** will be recruited by email invitation circulated on our behalf by the NRS Primary Care Network**.**

We plan to recruit these two groups of participants for one round of data collection only, anticipating that referral to long-COVID services will pick up in the coming months. However, should this assumption be incorrect, and referrals to services and this study remain low, we will include these two groups in subsequent rounds of data collection also.

***Informed consent***

Patients returning baseline measures will indicate their consent to taking part in the quantitative data collection by answering a question on the baseline measures paper or electronic form. Participants will self-select to take part in the focus group by leaving their name and contact details at the end of the survey or e-mailing the research team directly. The research fellow (RF) will then contact the participant by telephone or Microsoft Teams to discuss further and if relevant make arrangements for focus group participation. Consent to take part in the focus group will be provided verbally (audio recorded) during a separate telephone or Microsoft Teams call with the RF. A consent form with standard statements will be read to participants and they will have to agree to each statement in order to take part. Rehabilitation professionals and service leads/managers will provide verbal consent prior to commencing their interviews in the same manner.

Community-dwelling adults with long-covid will contact the research team using the details provided in invitation materials. Individuals will then be provided with study information and following discussion with the RF will provide verbal consent to be interviewed as described above.

GPs will likewise contact the research team following invitation emails sent by the NRS Primary Care Research Network, and will likewise provide verbal informed consent.

***Data collection***

***Patients:*** One online focus group will be conducted in each case study site during each round of data collection (n=4 focus groups of 8-10 participants each). Patients will be purposively sampled from those opting-in to a focus group, based on sampling criteria including age, gender, ethnicity, deprivation, and whether they completed rehabilitation or not. In the event that sufficient diversity is not achieved by this method, rehabilitation services will be asked to conduct additional targeted recruitment on our behalf. Focus groups will take place via Microsoft Teams and will each be conducted by 2 trained researchers using a topic guide. Focus groups will last no longer than 2-hours, will include adequate rest breaks, and will be recorded via Teams. To address issues of digital exclusion, a sample of participants who are interested in taking part but unable (or unwilling) to join an online focus group will be invited to provide their views via telephone interview or a short paper-based survey.

***Rehabilitation professionals and service leads/managers*** will take part in a 1-1 interview, conducted via Teams, supported by a topic guide and recorded in the same manner as the focus groups. From those volunteering to take part we will purposively recruit 3-5 professionals and 2 managers per site to be theoretically representative of staff at each site. We anticipate that the same staff will take part in each of the 4 rounds of data collection, unless there are changes in staffing, in which case additional recruitment will be conducted.

**Community dwelling adults and GPs** will take part in a single 1-1 interview, conducted by Teams, supported by a topic guide and recorded in the same manner as the focus groups. We will aim to recruit 3-5 community dwelling adults and 2-5 GPs at each site.

***Data processing***

Following data collection, the MP3 recordings will be downloaded by the RF and saved on secure servers at Robert Gordon University (RGU) and Stirling University (UoS) only accessible to the research team. The transcripts will be transcribed by GDPR-compliant transcribers and stored on the same secure servers.

***Data analysis***

Data will be analysed to articulate the context and mechanisms of action and relate these to observed (quantitative and qualitative) outcomes (CMO configurations). Each data source will be analysed individually in the first instance to reach separate conclusions. Data from meetings and interviews will be analysed using the Framework Approach (15) aided by NVivo (V12) software. For all qualitative datasets, 10% of transcripts will be coded independently by two RFs and the resulting codes discussed, refined and agreed as a final coding frame to be systematically applied to all transcripts. Following familiarisation with the data, a thematic framework will be developed and applied across the data set. Data will then be tabulated and conceptual maps used to make links between themes. Data will be used to construct case summaries at the level of the different modes of delivery and across different sites. All data for each mode of delivery and for each delivery site will be collected together and consistencies/ inconsistencies searched for. The aim of analysis at this stage is to identify the core barriers and facilitators and mechanisms of action within each mode of delivery, the detailed explanations for them and interactions between them.

### II. Outcome evaluation and cost-consequence analysis addressing outcomes 5-7

Outcomes data will be collected before and after people receive community rehabilitation in each location/model of service delivery to establish whether improvements are observable. Recruitment of up to 100 people per case study site will be conducted, from month 3 to month 18 of the study, by the staff delivering community rehabilitation in each local site. It is anticipated that each staff member (approximately 4-6 per team) will collect data on between 15-20 people over the 16-month period. The following patient demographic and clinical data will be collected by each team member for each consenting patient:

- Age
- Sex
- Ethnicity
- Occupation
- Social deprivation status in deciles (gathered using home postcode entered into https://simd.scot/#/simd2020/BTTTFTT/14/-2.7960/55.9548/
- Key presenting problems (e.g. musculoskeletal, fatigue, breathlessness, mental health)
- Co-morbidities (e.g. diabetes, hypertension, COPD)
- Smoking status
- Height & Weight (to estimate BMI)
- Duration of rehabilitation (date of first appointment to discharge)
- Number and duration of sessions
- Number and type of professionals seen

Patient outcomes will be collected at baseline immediately prior to first appointment, at the end of the last session of rehabilitation, and at 3-month follow-up. Baseline measures will be provided to patients with their appointment letter or at their first rehabilitation session (in person or by post if session is virtual), and they can complete the paper copy and return to the study team in freepost envelope or they can complete the measures online (Jisc online surveys) as a web address will also be provided.  Each participant will have a unique ID allocated by the site administration or clinical staff, which will be documented on the measures. Patients will be instructed to input this unique ID if they complete the measures online. Reminders will be sent by administration/clinical staff to follow-up individuals who do not submit completed measures. Regular communication between the study RFs and case study sites will facilitate reminders and sending discharge ad follow-up measures to participants.

#### *Outcome measures*

In keeping with the PHOSP study[1], we include the following outcomes:

[1]http://www.isrctn.com/ISRCTN10980107

***Primary Outcome***

**Quality of Life:** EQ-5D-5L utility index and visual analogue scale (VAS).

***Secondary Outcomes***

**Mental Health**

- Generalised Anxiety Disorder Assessment (GAD-7)
- Patient Health Questionnaire (PHQ-9)
- Post-Traumatic Stress Disorder Checklist for DSM-5 (PCL-5)

**Clinical symptoms**

- Dyspnoea12 Questionnaire
- FACIT – Fatigue Questionnaire

**Generic**

- Nottingham Extended Activities of Daily Living Scale
- General Practice Physical Activity Scale

**Covid Specific**

Post-COVID-19 Functional Status (PCFS)

***Service-related data***

In addition to the data collected from consenting patients, rehabilitation staff will collect the demographic and clinical data listed above (age, ethnicity, occupation etc.) for all long-covid referrals during the study period. This will allow us to determine the extent to which our sample is representative of the long-covid population in the four study sites.

***Data analysis***

Analysis of quantitative data will be address research aims: 3 (What are the outcomes of different community rehabilitation models for people experiencing long-covid?) and 4 (What are the costs associated with long-covid rehabilitation in the 4 selected cases?) Descriptive statistics will be tabulated for all outcomes at each time point, showing means and SDs. To describe representative changes in each outcome measured, generalized linear mixed models will be conducted. Mixed effects models will be used to account for the repeated measures nature of the data and appropriately address missing data without requirement for listwise deletion. Random intercepts and slopes will be included to quantify inter-individual variability in baseline measures and response to rehabilitation as well as general trends. Where suggested by the data and model checking processes, autoregressive models and non-normal link functions will be included to obtain more accurate parameter estimates and their associated standard errors. Addition of fixed effects into models will be included to investigate systematic differences associated with demographic factors (e.g. age, ethnicity, social deprivation status) and study sites where appropriate statistical power exists. Standard model building (e.g. modelling the effects of time [within-person fluctuation to within-person change over time], modelling the effects of predictors [time-invariant predictors to time-varying predictors in models of

within-person fluctuation and change]) and model checking (e.g. likelihood ratio tests and residual analyses) processes will be used throughout, with all analyses performed using the lme4 package in the statistical environment R.
The primary outcome is general health status measured via the EQ-5D-5L utility index and VAS. The EQ-5D-5L has been selected to differentiate smaller changes in health status and is less prone to ceiling effects (16). Based on NICE recommendations, quality-adjusted life-years will be calculated using tariffs specific to the EQ-5D-5L and will be mapped for reference-case analyses using the function developed by van Hout (17).
The costs associated with long-covid rehabilitation will be quantified through mean values for all outcomes presented for each study site with uncertainty estimated through boot-strapping procedures. Costs incurred at each site from the perspective of the NHS will be presented and will include estimated costs of the differing modes of delivery (digital, self-management and face to face). Unit costs will be obtained and calculated from Public Health Scotland's Cost Book for outpatient allied health professional clinics (R044X), summary of expenditure on community staff and supplies (R550), and community staff WTE and pay (SFR 8.1). ***Statistical Power:*** Estimates of statistical power were based on the primary outcomes and previous research of the minimum important difference for people with COPD. Multilevel mixed effects models were simulated in R using population parameters obtained from (16) and a correlation of 0.5 between repeated measures. With a two-sided alpha = 0.05, power to identify a common minimum important difference across the 4 study sites at both post-intervention and 3-month follow-up was estimated as 0.54, 0.74 and 0.87, for sample sizes of 60, 80 and 100, respectively.

***III. Synthesis of process, outcomes, and cost consequence analyses addressing research question 8***

Realist data synthesis will be undertaken. The findings from individual data sources (including participant outcome measures and data from interviews) will be presented in matrices that bring together key issues from the different analyses, to facilitate drawing overall conclusions about the mechanisms of change within each context, which components and mechanisms are most important, and how these might explain observed outcomes or unanticipated outcomes. A concurrent triangulation mixed methods design will be used (18), whereby both quantitative and qualitative data are considered simultaneously. Initially data from each source will be analysed separately (to reach separate conclusions) and then there will be cross-method analysis to reveal meta-inferences that will inform conclusions related to implementation models, context, mechanisms and outcomes (19).

**WP2:** "Dissemination for Implementation" workshops addressing research question 6. Two "Dissemination for Implementation" virtual workshops will be held: an interim one at month 12 and a final one at month 23. These online events will consist of information sharing and also an 'action planning' component for participants to reflect on how they can integrate study findings into their ongoing service delivery. These events will also act as 'data gathering' opportunities for the study, hence their inclusion as an objective: gathering data on potential opposition and barriers to implementation, how services make sense of and use our study findings and apply these to their own services. We expect the key people who lead and manage community rehabilitation services to attend and to leave with an action plan, and will use a targeted approach to promoting the workshops among the intended audience.

The workshops will be delivered according to a theoretically driven structure detailing: actions; theorised mechanisms of change; and expected outputs (See additional documentation). We have successfully used this theory driven workshop structure following both a Department of Health funded national consensus study (20) and an NIHR funded implementation study relating to implementation of a pelvic organ prolapse intervention (21). That workshop resulted in the study findings being adopted at a national level and the recommendations of the study and learning from the implementation day being implemented locally.

Workshop discussions will be audio-recorded (with permission) and/or notes will be taken by research team members on how participants translate study findings in relation to their local circumstances, any opposition or barriers, what level of local action planning is reached or reasons for non-action planning. These data will be qualitatively analysed using the Framework Approach, with specific attention to understanding levels of opposition and action, the discussion of barriers and whether the study findings influence the development of solutions to these barriers, whether study identified 'mechanisms' of change are adopted into local action plans and service delivery options, and which models of delivery seem more favourable for wider adoption and implementation across NHS Health Boards in Scotland.

### 6.0 Project Management

The study will be sponsored by RGU. It will be overseen by a Study Steering Committee (SSC) comprising of a Chair (independent academic researcher), one further independent academic researcher, a rehabilitation expert, two consumer representatives and the co-chief investigators (CIs). The SSC will meet quarterly. The CIs, co-applicants, and RFs will meet monthly as a Project Management Group (PMG) to consider the overall progress of the study and deal with any arising problems. Two consumer representatives will join the PMG quarterly. In addition to this, smaller sub-groups of members of the PMG will meet as required to ensure that individual elements of the study are progressing. Finally, KC and ED as Co-CIs will meet weekly with the study RFs about the day to day management of study activity. A full risk management plan will be developed prior to study commencement. This will be reviewed quarterly, during our PMG meeting. The project team will adhere to General Data Protection Regulation (GDPR) standards of data protection and data storage.

### 7.0 Ethical issues

The study will collect data from patient completed outcome measures, anonymous data on long covid rehabilitation service usage, and qualitative data from staff and people who have received long covid rehabilitation.

The populations being studied are adults (aged > 18 years) and NHS rehabilitation professionals. Potential participants will be either experiencing symptoms of long-covid, or be delivering a rehabilitation service within the context of a pandemic. Both situational contexts highlight the importance of clear participant information sharing about the nature and expectation of study participation and clear signposting that participation is entirely voluntary and can be withdrawn at any point without reason. Considerable consideration has been given to the data collection procedures to ensure that it is not burdensome on individuals or services. We have worked in close collaboration with the included services and with service users to co-construct research tools and agree recruitment and participation methods that are appropriate and proportional.

One of the key ethical concerns in this study is maintaining patient privacy, data management and data security. All study data will be stored on password protected and encrypted university servers (Sharepoint). Data will be re-identifiable in that participants will be allocated a code and coded data will be processed and analysed. The code sheet containing participants' identifiable details will be stored separately and only accessed where necessary (e.g. sending invites for interviews) by nominated members of the study team.

We considered at length and sought expert advice regarding participants potentially reporting clinically worrying scores on the outcome measures, especially GAD-7, PHQ-9 and PCL-5. As the research team will not access participants’ scores immediately, we are not in a position to intervene in a timely manner. Therefore, we have provided advice to participants about seeking help from their GP should they be concerned about their mental health after completing the measures (see participant information sheet), and we have also provided contact details for generic mental health support for all participants (also participant information sheet) as well as long-covid support. Focus group participants will also be provided with contact details for generic mental health support and long-covid support.

Having reviewed each of our study actions, we believe that risk of harm from this study is low. Notwithstanding this fact, the research team have established a study protocol to clarify communication and reporting channels if any element of the study methods proves to have significant unforeseen negative consequences. The protocol involves communicating initially with the relevant study site and forming an action plan to address the issue that has arisen. Where the unintended consequence could be relevant to other study sites, the PIs (KC, ED) will inform the pilot site leads of the situation and any suggested mitigating action that could be taken.

(14) long-covid survey

(15) Framework analysis

(13) Pawson R, Tilley N. Realistic evaluation: Sage; 1997.

(14) long-covid survey

(15) Framework analysis

(16) Nolan 2015. The EQ-5D-5: health status questionnaire in COPD: Validity responsiveness and minimum important difference.

(17) van Hout B et al. Interim scoring for the EQ-5D-5L: mapping the EQ-5D-5L to EQ-5D-3L value sets. *Value Health*. 2012 Jul-Aug;15(5):708-15**.**

(18) Creswell JW. Research Design: Qualitative, quantitative and mixed methods approaches. 2009 London: Sage.

(19) Teddlie CB, Tashakkori A. Foundations of Mixed Methods Research: Integrating Quantitative and Qualitative Approaches in the Social and Behavioral Sciences. 2009 London: Sage.

(20) Duncan, E et al. Consensus on items and quantities of clinical equipment required to deal with a mass casualties big bang incident: a national Delphi study. *BMC Emergency Med* 2020; 14(1): 5.

(21) Maxwell, M et al. PROPEL: implementation of an evidence based pelvic floor muscle training intervention for women with pelvic organ prolapse: a realist evaluation and outcomes study protocol. *BMC Health Services Research* 2017; 17(1):1-10.

### Appendix 1: Study Plan

| Year1 | | | | | | | | | | | | |
|---|---|---|---|---|---|---|---|---|---|---|---|---|
| **Month:** | **1** | **2** | **3** | **4** | **5** | **6** | **7** | **8** | **9** | **10** | **11** | **12** |
| | Site preparation & recruitment | | Qualitative Round 1 | | | | | Qualitative Round 2 | | | | |
| | | | | Qualitative data processing & analysis | | | | | | | | |
| | | | Quantitative data collection, processing & analysis | | | | | | | | | |
| | | | | Steering Committee | | | | Steering Committee | | | | Steering Committee |
| | | | | | | | | | | | | Workshop 1 |
| | PMG | PMG | PMG | PMG | PMG | PMG | PMG | PMG | PMG | PMG | PMG | PMG |
| **Year 2** | | | | | | | | | | | | |
| **Month:** | **1** | **2** | **3** | **4** | **5** | **6** | **7** | **8** | **9** | **10** | **11** | **12** |
| | Qualitative Round 3 | | | | | Qualitative Round 4 | | | | | | |
| | Qualitative data processing & analysis | | | | | | | | | | | |
| | Quantitative data collection, processing & analysis | | | | | | | Realist synthesis | | | | Reporting |
| | | | | | | | | | | | | Outputs |
| | | | | Steering Committee | | | | Steering Committee | | | | Steering Committee |
| | | | | | | | | | | | Workshop 2 | |
| | PMG | PMG | PMG | PMG | PMG | PMG | PMG | PMG | PMG | PMG | PMG | PMG |

PMG=Project Management Group

**Version Control**

| Date | Version | Changes |
|---|---|---|
| 23.03.21 | 1.0 | N/A |
| 07.03.22 | 2.0 | 1. Added objective 3 & 4 to explore community dwelling adults & GPs perceptions & experiences of long-covid rehabilitation<br>2. Updates participants, recruitment, consent, data collection for these additional objectives<br>3. Updated numbering of remaining objectives |
