## Supplementary File 2 for "Exploring the perceptions and experiences of community rehabilitation for Long COVID from the perspectives of Scottish General Practitioners’ and people living with Long COVID: a qualitative study" (from another version of this preprint: version 1, 2023-11-06)

### Consolidated criteria for reporting qualitative studies (COREQ): 32-item checklist

Please indicate in which section each item has been reported in your manuscript. If you do not feel an item applies to your manuscript, please enter N/A.

For further information about the COREQ guidelines, please see Tong *et al.*, 2017: https://doi.org/10.1093/intqhc/mzm042

| No. | Item | Description | Section # |
|---|---|---|---|
| **Domain 1: Research team and reflexivity** | | | |
| Personal characteristics | | | |
| *1.* | Interviewer/facilitator | Which author/s conducted the interview or focus group? | p7 |
| *2.* | Credentials | What were the researcher's credentials? *E.g. PhD, MD* | p8 |
| *3.* | Occupation | What was their occupation at the time of the study? | p8 |
| *4.* | Gender | Was the researcher male or female? | p8 |
| *5.* | Experience and training | What experience or training did the researcher have? | p7 |
| Relationship with participants | | | |
| *6.* | Relationship established | Was a relationship established prior to study commencement? | p8 |
| *7.* | Participant knowledge of the interviewer | What did the participants know about the researcher? *E.g. Personal goals, reasons for doing the research* | p8 |
| *8.* | Interviewer characteristics | What characteristics were reported about the interviewer/facilitator? *E.g. Bias, assumptions, reasons and interests in the research topic* | p8 |
| **Domain 2: Study design** | | | |
| Theoretical framework | | | |
| *9.* | Methodological orientation and theory | What methodological orientation was stated to underpin the study? *E.g. grounded theory, discourse analysis, ethnography, phenomenology, content analysis* | p6 |
| Participant selection | | | |
| *10.* | Sampling | How were participants selected? *E.g. purposive, convenience, consecutive, snowball* | p7 |
| *11.* | Method of approach | How were participants approached? *E.g. face-to-face, telephone, mail, email* | p7 |
| *12.* | Sample size | How many participants were in the study? | p9 |
| *13.* | Non-participation | How many people refused to participate or dropped out? What were the reasons for this? | p9 |
| Setting | | | |
| *14.* | Setting of data collection | Where was the data collected? *E.g. home, clinic, workplace* | p7-8 |
| *15.* | Presence of non-participants | Was anyone else present besides the participants and researchers? | p8 |

| | | | |
|---|---|---|---|
| *16.* | Description of sample | What are the important characteristics of the sample? *E.g. demographic data, date* | p10 (Table 1) |
| Data collection | | | |
| *17.* | Interview guide | Were questions, prompts, guides provided by the authors? Was it pilot tested? | p7 |
| *18.* | Repeat interviews | Were repeat interviews carried out? If yes, how many? | p7 |
| *19.* | Audio/visual recording | Did the research use audio or visual recording to collect the data? | p7 |
| *20.* | Field notes | Were field notes made during and/or after the interview or focus group? | p8 |
| *21.* | Duration | What was the duration of the interviews or focus group? | p8 |
| *22.* | Data saturation | Was data saturation discussed? | p28 |
| *23.* | Transcripts returned | Were transcripts returned to participants for comment and/or correction? | p8 |
| **Domain 3: analysis and findings** | | | |
| Data analysis | | | |
| *24.* | Number of data coders | How many data coders coded the data? | p8 |
| *25.* | Description of the coding tree | Did authors provide a description of the coding tree? | p112(Table 2) |
| *26.* | Derivation of themes | Were themes identified in advance or derived from the data? | p8 |
| *27.* | Software | What software, if applicable, was used to manage the data? | p8 |
| *28.* | Participant checking | Did participants provide feedback on the findings? | p9 |
| Reporting | | | |
| *29.* | Quotations presented | Were participant quotations presented to illustrate the themes / findings? Was each quotation identified? *E.g. Participant number* | p11-24 |
| *30.* | Data and findings consistent | Was there consistency between the data presented and the findings? | Yes |
| *31.* | Clarity of major themes | Were major themes clearly presented in the findings? | p11-24 & Table 2 |
| *32.* | Clarity of minor themes | Is there a description of diverse cases or discussion of minor themes? | p21 |

**When submitting your manuscript via the online submission form, please upload the completed checklist as a Figure/supplementary file.**

**If you would like this checklist to be included alongside your article, we ask that you upload the completed checklist to an online repository and include the guideline type, name of the repository, DOI and license in the *Data availability* section of your manuscript.**

Developed from: Allison Tong, Peter Sainsbury, Jonathan Craig, Consolidated criteria for reporting qualitative research (COREQ): a 32-item checklist for interviews and focus groups, International Journal for Quality in Health Care, Volume 19, Issue 6, December 2007, Pages 349–357, https://doi.org/10.1093/intqhc/mzm042
