## Supplementary File 3 for "Exploring the perceptions and experiences of community rehabilitation for Long COVID from the perspectives of Scottish General Practitioners’ and people living with Long COVID: a qualitative study" (from another version of this preprint: version 1, 2023-11-06)

LOng COvid Rehabilitation
In Scotland: an Evaluation

IRAS ID: 295672

**Study Title: Evaluating long-COVID rehabilitation in Scotland**

### Topic Guide for People with Long COVID

**Aim:** To explore community dwelling adults' perception and experiences of long-COVID and of attempting to access physical rehabilitation services

The interviews have a general purpose to obtain feedback from people with experience of long-COVID. The interviews will function in an open way to facilitate discussion of issues that are important to the individuals concerned. Therefore, the topic guide is deliberately broad and more specific areas for questioning will be driven by the interests of those taking part

#### General introduction

The researcher will introduce the purpose of the interview. The interviewee will provide their demographics (sex, age, employment status).

#### Interview questions

- Can you tell me a bit about your experience of long-COVID?
    - To put at ease, get some general background information, find out main symptoms/duration/progression

- Can you tell me about your experience of accessing healthcare for support with long-COVID?
    - Explore if/what/when/what happened/what helped or not
    - Explore if offered/accessed/tried to access rehabilitation services (e.g., community rehabilitation/physio/OT/other AHPs) and what the outcome was – if not tried to access why not
    - Explore if any barriers to accessing rehabilitation services
    - Explore if think rehabilitation services could be helpful for them or others with long-covid
    - Explore if they consider themselves to have unmet needs regarding management of long-COVID

- Is there anything else you would like to tell me about your experience of long-COVID or accessing healthcare services that you think would be useful to our study?

LOng COvid Rehabilitation
In Scotland: an Evaluation

IRAS ID: 295672

### Topic Guide for GPs

**Aim:** To explore GPs experiences of managing people with long-covid, in particular people who have symptoms that may benefit from rehabilitation from AHPs (e.g., physiotherapists, occupational therapists, dietitians, speech and language therapists), and their experiences of long-covid rehabilitation services in their area

The interviews have a general purpose to obtain feedback from those involved in managing people with long-covid. The interviews will function in an open way to facilitate discussion of issues that are important to the individuals concerned. Therefore, the topic guide is deliberately broad and more specific areas for questioning will be driven by the interests of those taking part

#### General introduction

The researcher will introduce the purpose of the interview. The GP will introduce their role, years' experience as GP, and general experience of long-COVID.

#### Interview questions

- Do you have many patients presenting with long-COVID?
  - Explore numbers/how being "diagnosed"/presenting problems
- How are you managing patients with long-COVID?
  - Explore whether individualised or whether any trends for what patients need
  - Explore self-management/onward referral (where to)
  - Explore if perceived need for rehabilitation (e.g., for reduced exercise tolerance, breathlessness, diet, swallowing etc.)
    - If yes next question
    - If no explore why not?
- What are your perceptions of the value of services such as community rehabilitation for people presenting with long-covid symptoms?
- What community rehabilitation services are available for long-COVID patients in your practice to be referred to?
  - Explore whether aware of CRTs or individual professions (e.g., physio/OT) who may be taking referrals
  - Are there any barriers to referring patients to community rehabilitation?
- Anything else you would like to say about managing patients with long-COVID that might be of use to this study?
